# The preparedness and response to COVID-19 in a quaternary Intensive Care Unit in Australia: perspectives and insights from frontline critical care clinicians

**DOI:** 10.1101/2021.11.05.21265998

**Authors:** Krishnaswamy Sundararajan, Peng Bi, Adriana Milazzo, Alexis Poole, Benjamin Reddi, Mohammad Afzal Mahmood

**Affiliations:** Staff Specialist, Intensive Care Unit, Royal Adelaide Hospital and Discipline of Acute Care Medicine, The University of Adelaide; Professor, School of Public Health, The University of Adelaide; Senior Lecturer, School of Public Health, The University of Adelaide; Critical Care Registered Nurse, Intensive Care Unit, Royal Adelaide Hospital and Discipline of Acute Care Medicine, The University of Adelaide; Senior Lecturer, School of Public Health, The University of Adelaide and Faculty of Medicine, Universitas Airlangga, Indonesia

**Author notes:** Corresponding author: Dr Krishnaswamy Sundararajan, Head of Unit, Level 4, Intensive Care Unit, Royal Adelaide Hospital, Port Road, Adelaide, South Australia-5000. Funding source(s): None. Declarations: None. Contributions KS was involved with the conceptualisation of the study, literature review, ethics application, conduct of the study and investigation, methodology and writing the original draft. PB was involved with the methodology, data analysis editing the manuscript. AM was involved with the writing review. AP and BR were involved with the ethics submission, methodology, data analysis and writing review. MAM was involved with literature review, methodology, data analysis and editing the manuscript. Patient or substitute decision maker’s consent for publication: Not applicable. Data sharing statement: Data has been stored with the University of Adelaide repository and will be provided on request.

**Keywords:** COVID-19, Intensive Care Unit, Preparedness, Response, Hospital Management, Communication, Resilience, Command and Control

## Abstract

**Objectives:** This study was conducted to explore the perspectives and opinions of Intensive Care Unit (ICU) nurses and doctors at a COVID-19 designated pandemic hospital concerning the preparedness and response to COVID-19 and to consolidate the lessons learnt for crisis/disaster management in the future.

**Design:** A qualitative study using in-depth interviews (IDIs) and focus group discussions (FGDs). Purposeful sampling was conducted to identify participants. A semi-structured guide was utilised to facilitate in-depth interviews with individual participants. Two focus group discussions were conducted, one with the ICU doctors and another with the ICU nurses. Thematic analysis identified themes and subthemes informing about the level of preparedness, response measures, processes, and factors that were either facilitators or those that triggered challenges.

**Setting:** ICU in a quaternary referral centre affiliated to a university teaching COVID-19 designated pandemic hospital, in Adelaide, South Australia.

**Participants:** The participants included eight ICU doctors and eight ICU nurses for the in-depth interviews. Another sixteen clinicians participated in focus group discussions.

**Results:** The study identified six themes relevant to preparedness for, and responses to, COVID-19. The themes included: (1) Staff competence and planning, (2) Information transfer and communication, (3) Education and skills for the safe use of PPE, (4) Team dynamics and clinical practice, (5) leadership, and (6) Managing End-of life situations and expectations of caregivers.

**Conclusion:** Findings highlight that preparedness and response to the COVID-19 crisis were proportionate to the situation’s gravity. More enablers than barriers were identified. However, opportunities for improvement were recognised in the domains of planning, logistics, self-sufficiency with equipment, operational and strategic oversight, communication, and managing end-of-life care.

**ARTICLE SUMMARY:** *Strengths and limitations of this study:*

- This is the first study that provided insights about clinicians’ perspectives and viewpoints to preparing and responding to COVID-19 in Australia.
- The study used qualitative methodological framework allowing participants to provide in-depth accounts of processes and enabling factors and barriers.
- Our study provides information on issues that needs to be addressed from a critical care viewpoint and interventions that were effective and efficient
- This is a single-center study in a developed country where experience is vastly different from other centers with higher demand and fewer resources
- We acknowledge the potential for selection bias because of the qualitative design

## INTRODUCTION

The COVID-19 pandemic caused by the pathogen severe acute respiratory syndrome coronavirus 2 (SARS-CoV-2) presents an ongoing public health threat with the possibility of a massive influx of critically ill patients into a system with limited capacity despite the recent rollout of vaccination programs. Sharing guidance [1] between various hospitals at a national level could promote effective planning as the burden on ICU resources in the context of COVID-19 is likely to be significant. Many features of the current crisis make sustained surge capacity [2] a concern. In these circumstances, a command-and-control model is often instituted as a strategy to delineate roles and responsibilities, ensure appropriate allocation of resources, and maintain clinical service delivery, including business as usual. This model’s utility in facilitating a coordinated response to COVID-19 pandemic in an ICU has not been fully explored.

The short, medium, and long-term consequences of the COVID-19 pandemic are still very uncertain. Therefore, clinicians in the field of intensive care medicine have relied on existing evidence-based policies, procedures and guidelines promulgated by professional bodies.[3] A respected source of information [4] in the Australian context has been the Australia and New Zealand Intensive Care Society guidelines. These guidelines along with government directives [5] cover the practical aspects concerning the use of personal protective equipment (PPE) and isolation rooms. The uptake and compliance with these guidelines and directives related to COVID-19 among frontline clinicians working in an ICU, however, have not been fully elucidated.

Further, managing end-of-life care for patients with COVID-19 has been challenging. Intensive care resources are used quite commonly in managing terminal hospitalisations and end-of-life care in the developed world [6]. Coupled with the emerging body of evidence in relation to prognostication and fatality rates [7, 8] around COVID-19, the presumed increase in the infectivity of patients at the end of life compounds the challenges of providing good palliative care [9]. In Australasian ICUs, the standards in relation to consumer engagement and comprehensive care including end-of-life care highlight the importance of dealing with patients and their substitute decision makers with empathy and compassion [10-12].

### Rationale

The perspectives of those directly involved in the care of COVID-19 patients in an ICU and the level of preparedness in terms of competence and infrastructure for providing care for patients with a highly transmissible disease are unknown. By ‘competence’ we mean the skills and knowledge needed for managing COVID-19 in a critically ill patient. In the same vein, the processes by which staff adapt and prevail in those circumstances, and barriers and enablers in communicating effectively in a pandemic, particularly within the governance of a crisis management framework are not well understood. In addition, given the dynamic nature of the pandemic, the practical relevance and utility of simulation sessions in educating and empowering frontline clinicians in undertaking high-risk procedures is not clear. Moreover, managing end-of-life situations, during COVID-19 in an Australian context, is not well understood. This study addresses these knowledge gaps.

### Objectives

This study explored the perspectives and opinions of ICU nurses and doctors in relation to the preparedness and response to COVID-19 and identify enablers and barriers to consolidate knowledge for future crisis/disaster management in a quaternary referral ICU in a hospital designated for pandemics.

## METHODOLOGY

### Study design

Using phenomenology [13,14], we adopted a qualitative approach [15,16] to explore the participants’ perspectives and viewpoints, using in-depth interviews (IDIs) as well as focus-group discussions (FGDs). The structure and the topics explored were the same in the IDIs and FGDs. The study was designed to allow exploration of the lived experience of critical-care professionals, not only in terms of preparedness but also in terms of their response to the pandemic as an interpretive process and this aligned with hermeneutic (interpretive) phenomenological approach [17]. The guidelines for standard reporting of qualitative research were upheld for that methodology [18]

### Setting

The study was conducted in a 48-bedded ICU of an 800-bedded quaternary hospital in Australia between August and October 2020, where critically ill patients with COVID-19 were being managed. The COVID status matrix illustrating the horizontally and vertically integrated, command and control structure named COSTAT was used for crisis management and around the time of this crisis, the organisation reached the maximum staging of COSTAT 4. (See supplemental file) The COSTAT matrix reflected the fact that while critical decisions were centrally regulated at the organisational level (control: vertical); decisions were also made at various levels within the ICU (command: horizontal)

### Study participants

Purposeful sampling was undertaken to recruit participants from the medical and nursing divisions of the ICU. These frontline clinicians were approached at ICU staff forums which provided the opportunity to apprise the clinicians about the study and solicit their participation. For those who expressed interest, further sessions were arranged at a mutually convenient time to discuss the study in more detail, explaining the aims, methods, ethical considerations, potential risks, and a recourse to withdrawal of consent at any stage. A total of 18 respondents were invited for the in-depth interviews, of whom 16 accepted to be interviewed. For the focus group discussions, 25 staff members were invited, of whom 16 were able to participate. There was no remuneration for those who participated. The interviews were conducted between 10th August 2020 and 27th October 2020. The discussion topics were generated based on a pilot run and around the practical issues that mattered the most to all staff around that time

### Governance

This study was approved by the Central Adelaide Local Health Network, Human Research Ethics Committee (Reference number 13363).

IDIs and FGDs were conducted after obtaining fully informed, written, and signed consent from each study participant. We ensured the participants’ confidentiality by allocating a unique study number to their interviews, and we de-identified participants’ details for both in-depth interviews and focus-group discussions.

### Patient and public involvement

No patients, substitute decision makers or members of public were involved in this study.

### Data curation and Reflexivity

As data curator, the first author led the process of data collection for the face-to-face IDIs and FGDs. Lived experience along with the views and perceptions of frontline clinicians at the coalface was best elucidated by the IDIs. The FGDs complemented the interviews’ findings through their ability to corral team members, cross-validate their opinions and perceptions and consolidate their views. All IDIs and FGDs, with permission from the participants, were recorded using an audio software on smartphones and transcribed verbatim for analysis. The participants were provided with the transcripts and the opportunity to clarify responses before analysis. No participant withdrew from the study.

### Data analysis

We used an archive (Box®, a web-based document-sharing platform with password protection) to store the transcribed versions of the audio-recorded interviews, which facilitated sharing information between investigators and the subsequent analysis. Thematic analysis, according to the ‘framework approach’ described by Spencer and Ritchie et al. [19] and adopted by Strickland and Hackett [20], was employed. The data analytics using the above-mentioned approach was led by the first author and the data analysis scheme* is illustrated in Figure 1

**Figure 1.**
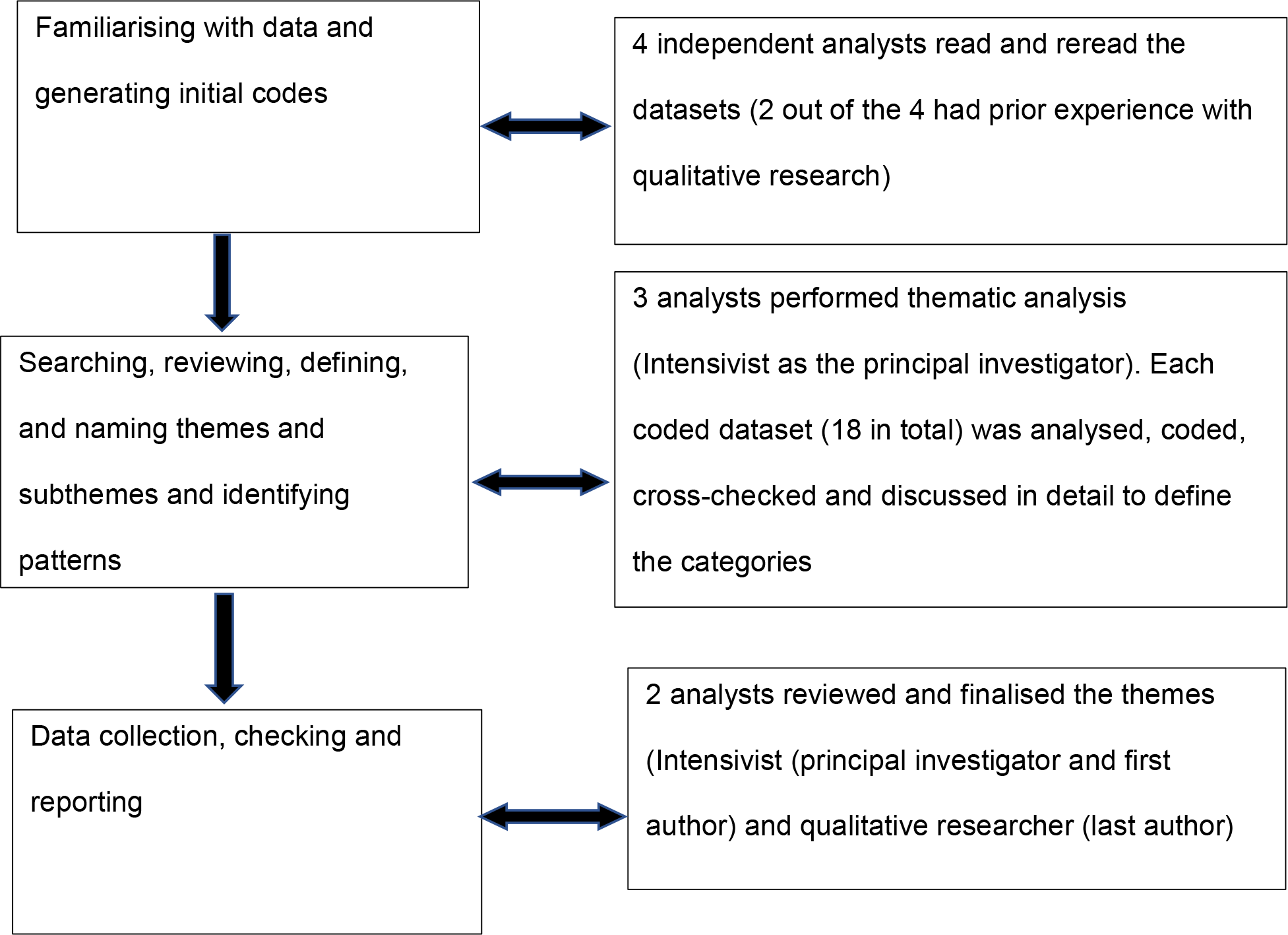
Data analysis scheme

### Fidelity

We addressed the study’s trustworthiness by enhancing its internal and external validity and its credibility, transferability, and reliability. To enhance credibility, we used a well-validated research methodology (qualitative descriptive) [21-22] and combination of methods (IDIs and FGDs) based on purposeful sampling, using an interview guide that used probes to facilitate the IDIs and FGDs

All interviews were audio recorded and transcribed professionally with ethics approval; this approach generated high-quality transcripts that included direct quotations in 18 datasets (16 individual interviews and two focus groups).

From these datasets, we identified similarities in the codes, themes, and sub-themes, thereby ensuring the data’s veracity. The participants concurred that the results reflected their true viewpoints, thus strengthening our study’s internal validity and rigour. The coding was initially done by the principal investigator, which was overseen by the last author. The coding undertaken by the manual process was crosschecked with the coding obtained from the Nvivo 12 software to enhance the authenticity of the codes and the reliability of the study.

For generalisability, we have outlined the research settings and the participants i.e., a quaternary referral ICU at a university teaching hospital with trained critical-care nurses and accredited doctors under the auspices of the College of Intensive Care Medicine of Australia and New Zealand, as the finding might strike a chord with similar centres across the world.

## RESULTS

Thirty-two participants took part in sixteen IDIs and two FGDs (eight participants in each group). As data saturation was achieved and no new barriers and enablers were identified, after sixteen IDIs and two FGDs; no further interviews were undertaken.

For the IDIs (n=16), participants comprised seven (44%) women and nine (56%) men, which included eight (50%) intensivists and eight (50%) critical care trained nurses. For the FGDs (n=16), ten (63%) women and six (37%) men who were not involved in the interviews participated, which included eight (50%) intensivists and eight (50%) critical care trained nurses. All participants were involved in direct care of COVID-19 patients. The demographic data has been summarised in Figure 2. The interview guide for the in-depth interviews and the moderator guide for the focus group discussions have been added to the supplemental file. We identified six themes and twenty-four sub-themes concerning the preparedness for, and responses to, COVID-19. The themes and sub-themes are summarised in Table 2.

**Table 1.**
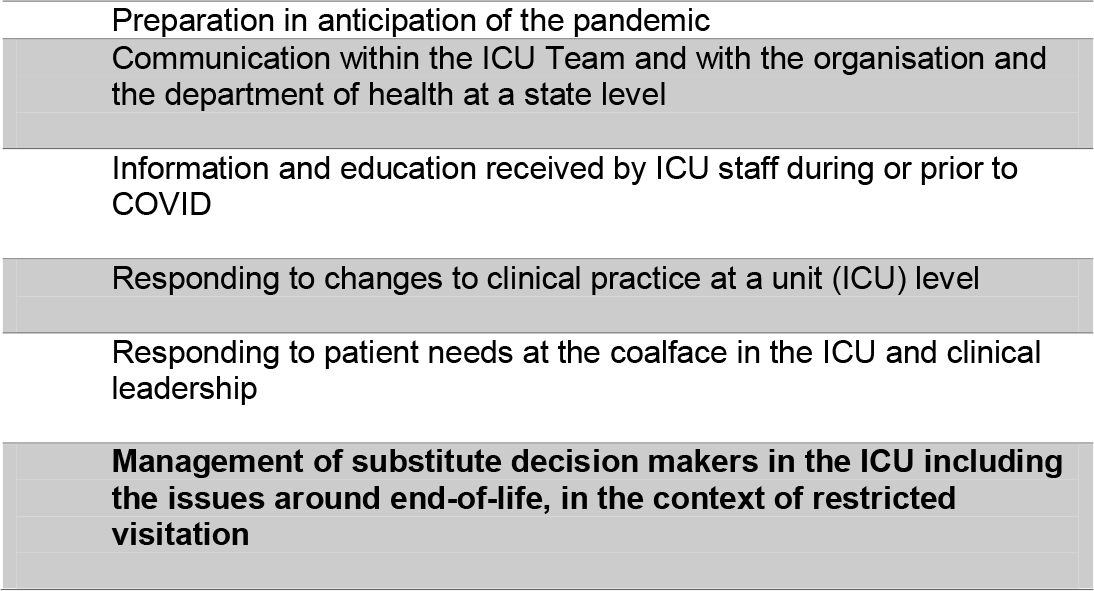
Structure for in-depth interviews and focus group discussions

| Structure for in-depth interviews and focus group discussions |
| --- |
| Preparation in anticipation of the pandemic |
| Communication within the ICU Team and with the organisation and the department of health at a state level |
| Information and education received by ICU staff during or prior to COVID |
| Responding to changes to clinical practice at a unit (ICU) level |
| Responding to patient needs at the coalface in the ICU and clinical leadership |
| Management of substitute decision makers in the ICU including the issues around end-of-life, in the context of restricted visitation |

**Table 2:**
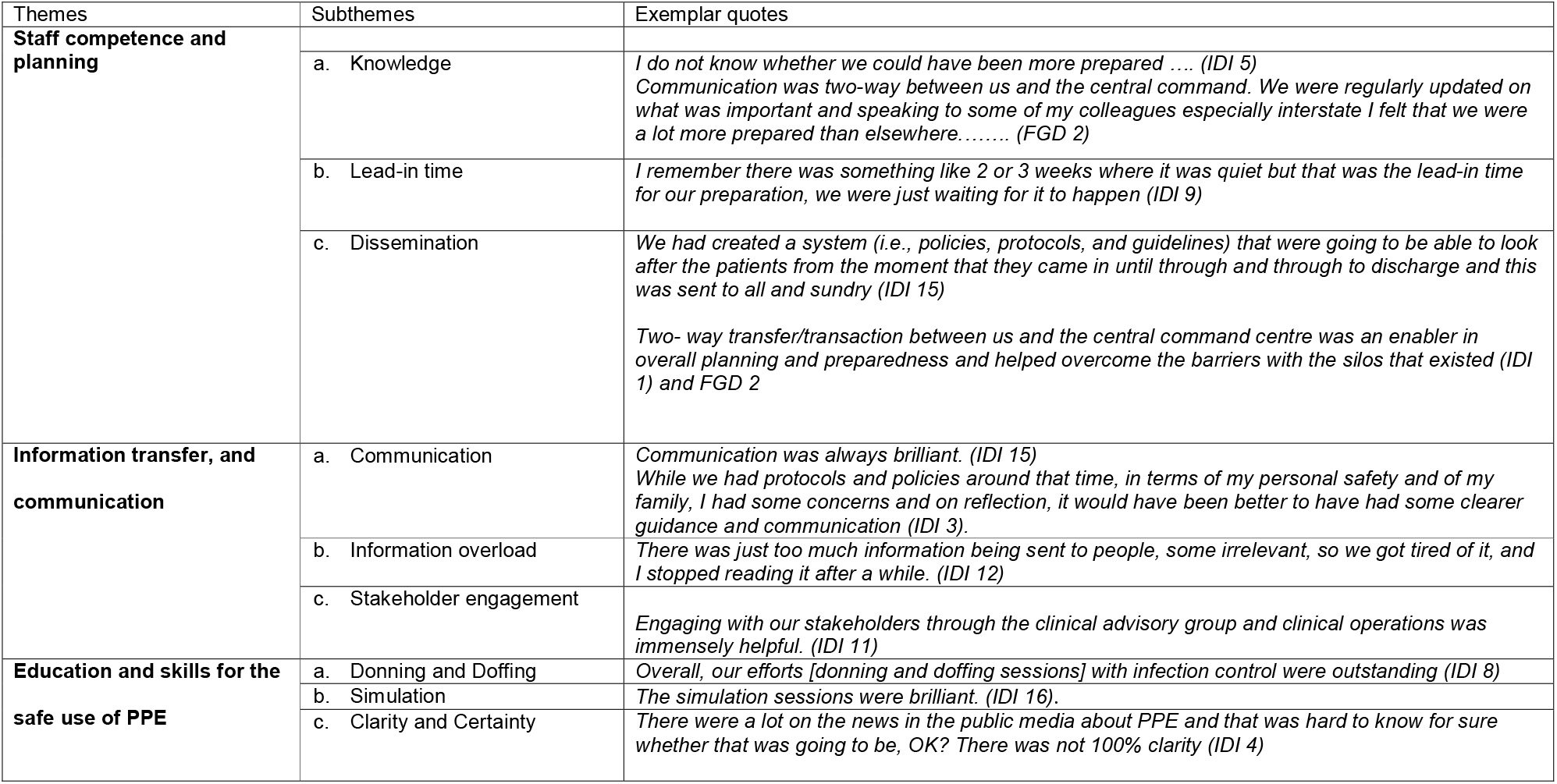
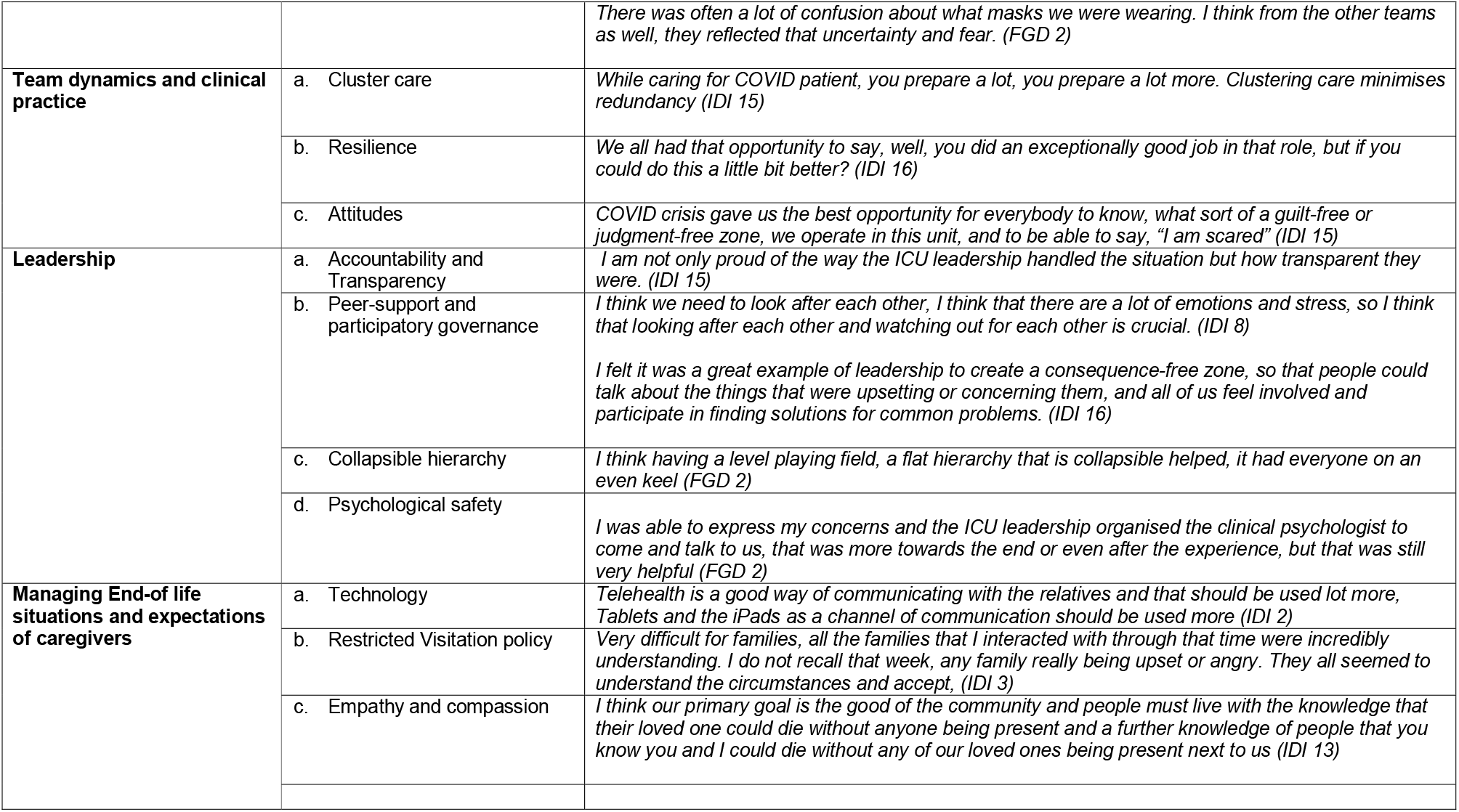
Themes, Sub-themes, and exemplar quotes written verbatim from the clinicians illustrating the enablers and barriers.

| Themes | Subthemes | Exemplar quotes |
| --- | --- | --- |
| **Staff competence and planning** |  |  |
|  | a. Knowledge | *I do not know whether we could have been more prepared .... (IDI 5)*<br>*Communication was two-way between us and the central command. We were regularly updated on what was important and speaking to some of my colleagues especially interstate I felt that we were a lot more prepared than elsewhere..... (FGD 2)* |
|  | b. Lead-in time | *I remember there was something like 2 or 3 weeks where it was quiet but that was the lead-in time for our preparation, we were just waiting for it to happen (IDI 9)* |
|  | c. Dissemination | *We had created a system (i.e., policies, protocols, and guidelines) that were going to be able to look after the patients from the moment that they came in until through and through to discharge and this was sent to all and sundry (IDI 15)*<br>*Two- way transfer/transaction between us and the central command centre was an enabler in overall planning and preparedness and helped overcome the barriers with the silos that existed (IDI 1) and FGD 2* |
| **Information transfer, and communication** | a. Communication | *Communication was always brilliant. (IDI 15)*<br>*While we had protocols and policies around that time, in terms of my personal safety and of my family, I had some concerns and on reflection, it would have been better to have had some clearer guidance and communication (IDI 3).* |
|  | b. Information overload | *There was just too much information being sent to people, some irrelevant, so we got tired of it, and I stopped reading it after a while. (IDI 12)* |
|  | c. Stakeholder engagement | *Engaging with our stakeholders through the clinical advisory group and clinical operations was immensely helpful. (IDI 11)* |
| **Education and skills for the safe use of PPE** | a. Donning and Doffing | *Overall, our efforts [donning and doffing sessions] with infection control were outstanding (IDI 8)* |
|  | b. Simulation | *The simulation sessions were brilliant. (IDI 16).* |
|  | c. Clarity and Certainty | *There were a lot on the news in the public media about PPE and that was hard to know for sure whether that was going to be, OK? There was not 100% clarity (IDI 4)* |
|  |  | *There was often a lot of confusion about what masks we were wearing. I think from the other teams as well, they reflected that uncertainty and fear. (FGD 2)* |
| **Team dynamics and clinical practice** | a. Cluster care | *While caring for COVID patient, you prepare a lot, you prepare a lot more. Clustering care minimises redundancy (IDI 15)* |
|  | b. Resilience | *We all had that opportunity to say, well, you did an exceptionally good job in that role, but if you could do this a little bit better? (IDI 16)* |
|  | c. Attitudes | *COVID crisis gave us the best opportunity for everybody to know, what sort of a guilt-free or judgment-free zone, we operate in this unit, and to be able to say, "I am scared" (IDI 15)* |
| **Leadership** | a. Accountability and Transparency | *I am not only proud of the way the ICU leadership handled the situation but how transparent they were. (IDI 15)* |
|  | b. Peer-support and participatory governance | *I think we need to look after each other, I think that there are a lot of emotions and stress, so I think that looking after each other and watching out for each other is crucial. (IDI 8)*<br>*I felt it was a great example of leadership to create a consequence-free zone, so that people could talk about the things that were upsetting or concerning them, and all of us feel involved and participate in finding solutions for common problems. (IDI 16)* |
|  | c. Collapsible hierarchy | *I think having a level playing field, a flat hierarchy that is collapsible helped, it had everyone on an even keel (FGD 2)* |
|  | d. Psychological safety | *I was able to express my concerns and the ICU leadership organised the clinical psychologist to come and talk to us, that was more towards the end or even after the experience, but that was still very helpful (FGD 2)* |
| **Managing End-of life situations and expectations of caregivers** | a. Technology | *Telehealth is a good way of communicating with the relatives and that should be used lot more, Tablets and the iPads as a channel of communication should be used more (IDI 2)* |
|  | b. Restricted Visitation policy | *Very difficult for families, all the families that I interacted with through that time were incredibly understanding. I do not recall that week, any family really being upset or angry. They all seemed to understand the circumstances and accept, (IDI 3)* |
|  | c. Empathy and compassion | *I think our primary goal is the good of the community and people must live with the knowledge that their loved one could die without anyone being present and a further knowledge of people that you know you and I could die without any of our loved ones being present next to us (IDI 13)* |

**Figure 2.**
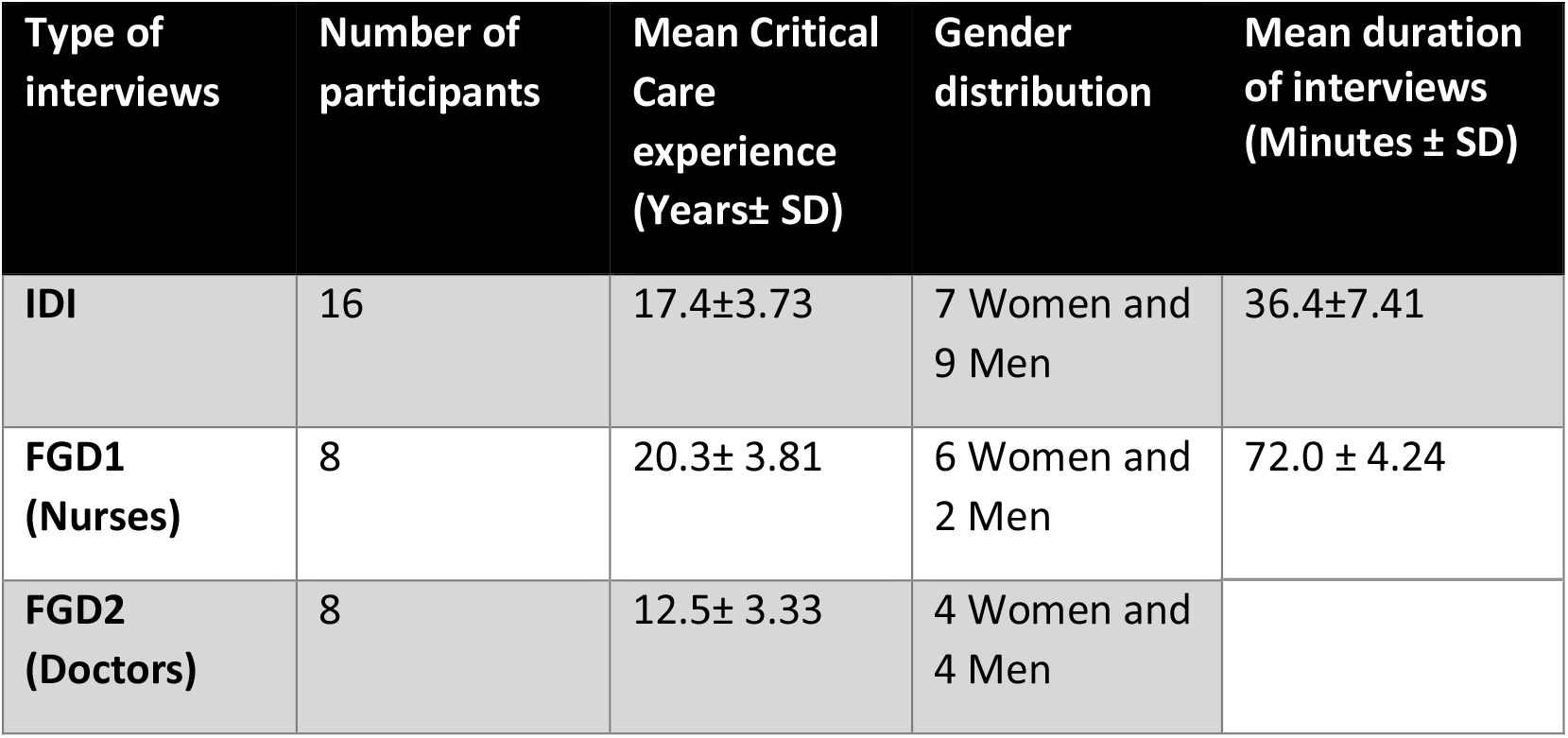
Demographic data on participants

### Staff competence and planning

This theme describes the contrary perspectives of ICU nurses and doctors on staff competence and planning. The concept of competency includes both the knowledge (what to do) and the skills (how to do it) necessary to be prepared and respond effectively and efficiently to COVID-19.

> ***“I felt like the ICU team were well prepared and that goes to all the different aspects of the preparation*.*” ……. (IDI 4)***
>
> ***“The amount of work that was done in a very short span of time was commendable” ………***..***(IDI 9)***

Some healthcare providers compared what was happening at the national level and opined that the organisation was better prepared compared with other centres within Australia. The lead-in time (i.e., the time lag between the COVID-19 outbreak overseas, and the subsequent arrival of the cruise ships on Australian shores) was identified as a stroke of good fortune by most participants (*n* = 12, 75%).

> ***“It did not seem like we pushed hard until COVID in Italy exploded, we picked up after Italy and it was then we put the foot on the accelerator” (IDI 9)***

Planning and lead-in time were also key factors that influenced the ability to provide staff with knowledge and access to important information about the protocols, policies, and guidelines pertinent to the management of COVID-19.

There was general agreement among the participants that the most recent and updated version of the policies, protocols and guidelines concerning the management of patients with COVID-19 were easily available. Perceived barriers in the planning phase were related to the silos that existed with different levels of procurement of equipment. However, the command-and-control structure was recognised as an enabler in overcoming those barriers.

> ***You suddenly find out all the things (i*.*e***., ***PPE) that are not produced locally, and we had to procure them at short notice. (IDI 5)***

### Information transfer, and communication

Participants expressed divergent views on the quality and quantity of communication, the nature of the transfer of information and, more importantly, the ‘dose’ of information, as information overload emerged as one of the themes. The perceived shortcomings in terms of inconsistency of communication were a source of healthcare providers’ frustrations. Quality refers to the attributes of timely communication of evidence-based information, while quantity describes the volume of evidence-based data.

On the other hand, communication between doctors and nurses (horizontal information flow) and between the command-and-control structure and the frontline workers (vertical information flow) was optimal in dose (quantity) at several stages of preparation and response. The command-and-control structure used an incident management framework [23] that linked the executive medical administration, heads of units and divisional leads to the various clinical programs (i.e., specialities) of the hospital (i.e., command) and the health department at the state level (i.e., control). This framework also integrated various aspects of intelligence gathering, planning, operations, logistics, public information, investigations, and recovery. Innovative use of technology, such as web-based document-sharing platform (Dropbox®), for instant sharing of information; WhatsApp notifications for urgent issues; and Zoom meetings for virtual conferences, meant that it was easy to access information throughout the day.

***“I think our consultant group were very supportive of the nursing team and were approachable. Had no problems in communication “(IDI16)***

A few participants made the distinction between information and fact. Further, sharing value sets (collegiality and patient centeredness) and vulnerabilities (acknowledging our fears) was recognised as a vital element of having meaningful and deep engagement with key stakeholders. Our key stakeholders involved the medical personnel from the emergency department, infectious diseases, general medicine, anaesthesia, surgical specialties, and the nursing & allied health staff from the above-mentioned craft groups.

***“And there was a lot of information and not much fact…. (IDI 7)***

### Education and skills for the safe use of PPE

The sub-themes revolved around donning-and-doffing sessions, educational lessons on the safe use of PPE and simulation exercises, including mock drills on critical scenarios. Participants were unanimous in acknowledging the utility and validity of the simulation sessions, which were very helpful, not just at an individual level in terms of improving their skills, confidence, and competence but also at a unit level by improving the quality of clinical service delivery, throughput and operational efficiency and effectiveness.

***“I think that simulation has really helped me in terms of practising life sustaining skills and then applying them in practice. (IDI 8)***

### Team dynamics and clinical practice

‘Cluster care’ is the mechanism by which multiple patient-focused tasks are integrated and doctors and nurses collaborate to minimise interruptions to the time spent by nurses in patient cubicles; this may involve delegating tasks to doctors, which otherwise would not have occurred, the importance of cluster care and team dynamics came to light during both in-depth interviews and focus groups.

Simultaneously, the respondents acknowledged that, while having agreement on policies, protocols and guidelines, minimised unwarranted clinical-practice variation, the risk of such variation was not nullified completely. Teamwork and cohesiveness were essential drivers in mitigating unwarranted clinical-practice variation. Cluster care emerged as a new phenomenon and is likely to be used more frequently because it was perceived by the informants as an effective and efficient mechanism of addressing redundancy and non-value-added work.

***Clustering care is the way forward as it is efficient and effective (FGD 2)***

***Consistency, cohesiveness, transparency, and teamwork with a pipeline to escalate issues would help minimise clinical practice variation (FGD 2)***

### Leadership

The participants were unanimous in their views that the leadership was decisive and accountable during pandemic management. However, respondents did highlight some inconsistencies and incongruence in delivering unambiguous take-home messages that, in hindsight, stemmed from the difficulties in reconciling information from various sources. A flat or collapsible hierarchy, at the unit where this study was conducted, that emboldens the tenets of good corporate citizenry, peer support and psychological safety was highlighted as one of the critical enablers. The participants had divergent views of the lines of communication in terms of who was responsible, who was accountable, who was to be consulted and who was to be informed, and it was suggested by the respondents that one of the lessons from this experience would be to streamline the lines of reporting and escalation pathways.

***I thought that certainly from a hospital level, the leadership at a clinical and managerial level was strong and sensible and more importantly visible and accountable (FGD 2)***

### Managing end-of-life situations and expectations of caregivers

All participants were of the view that managing the end-of-life situation in COVID-19 patients was the most challenging experience in their professional lives. The group was unanimous that technology was beneficial. Had it not been for video-calling devices (i.e., iPads/tablets), communicating with families would have been impossible, and the situation would have been a lot more traumatic for all concerned.

> ***I think for the families whilst distressing on one hand to see how their loved one is going; it is kind of comforting as well to know that there is somebody with them. (IDI 12)***

## DISCUSSION

This is the first study conducted to explore the perspectives, opinions and proficiencies of staff managing critically ill patients in an ICU of a hospital designated for managing COVID-19 in Australia. It addresses the knowledge gap in how frontline clinicians adapt to these circumstances. This study identified more facilitators than impediments in providing care to those patients. Findings inform that with effective communication and teamwork it is possible to provide effective care to patients and support for staff despite the huge challenges posed by a pandemic. The facilitators were identified in the creative use of technology in galvanising the workforce, dealing with difficult family situations, and engaging with families. Use of simulation technology was also a primary facilitator, as were shared values among colleagues and a spirit of collegiality, which helped practitioners to prevail over the barriers. The barriers revolved around information overload and inconsistent messaging. The lack of clarity on PPE use and its perceived non-availability point to concerns with pandemic preparedness and the distribution of vital resources to those on the frontlines.

Overall, the clinician’s perspectives on staff competence and planning were favourable, notwithstanding the divergence in opinions regarding planning at various stages of the pandemic. While it may be tempting to attribute the competence and planning to overall preparedness, the importance of the lead-in time before the first wave hit the Australian health system cannot be overstated.

The lead in time allowed the organisational machinery to build capacity, marshal resources and, more importantly, allow people to get into the right headspace to stand and deliver. As noted by the previous research [24], we also found that the leadership in this unit facilitated trust and empowerment and that contributed to hospital’s readiness and resilience the teamwork and stakeholder engagement were an enabler in planning and execution. This finding is consistent with the evidence that organisations that believe in a ‘people first’ policy foster resilient healthcare [25] and have better outcomes and performance. Technology and health informatics [26] were enablers in disseminating information and keeping staff members updated on current guidelines. There was good uptake using the Dropbox platform, video conferencing and virtual meeting facilities and tele-counselling to provide emotional support to caregivers of patients with COVID-19. This finding mirrors those of other studies [26,27] that have identified technology as a valuable resource in crisis management and in reorganising clinical workflows to make them more efficient and robust.

Information overload was one of the main concerns that emerged from this study. This resonates with the experiences of other frontline workers across the globe [28]. Information asymmetry and discordance were the other parallel themes which aligns with the issues highlighted by other organisations in similar situations [29]. The COVID-19 pandemic has also been described as the ‘infodemic’ [29]. Technology cuts both ways; however, while the appropriate use of technology can make it easier to share useful information, the unintended consequence of attempting to share information quickly is the perception of mixed messaging when specifics crystallise over time.

An important finding relates to skill-enhancement methods, highlighted by the respondents’ concern with the education and skills needed to use PPE. The donning-and-doffing sessions [30] and simulation exercises (mock drills) [31] provided the clinicians with real-life experience; the practice was unanimously labelled as a very valuable exercise. These findings mirror the emerging body of literature [30,31] on the pandemic. PPE has been a pivotal issue [32] in this pandemic owing to several concerns; for the first time in recent memory, clinicians were stressed about the risk not only of asymptomatic transmission [33] of the virus, but also of the impact, inadequate or suboptimal PPE could have on their patients, close family members [34] and the community. Numerous healthcare workers have either acquired the disease, suffered moral distress and burnout, or died in the line of duty.[35] Participants expressed a view that, while efforts to mobilise resources, undertake fit-testing and communicate effectively was better than at most centres,[36] the clarity of information and efficiency in sharing intelligence concerning PPE was lacking, the National COVID-19 Clinical Evidence Taskforce only recently have been provided with resources to provide independent evidence based guidelines on various PPE to support frontline workers and this was identified as an opportunity for improvement.

Work dynamics, team culture and peer support emerged as central themes. The core of them was resilience. According to our participants, the combination of having foresight on how the pandemic would unravel, its implications on clinical practice, and the need to modify workflows in response to contingencies. The other aspects of resilience that were tested were the ability to cope under pressure, disallowing a bad situation to escalate; and the tenacity to recover without suffering the consequences of burnout. This experience was sentinel amongst the clinicians and mirrors the emerging research on resilience in organisations.[37] Psychological support was identified as an enabler, but the majority felt it was offered late in the process and could have been considered earlier. [37,38] Resolute and decisive leadership is a key requirement [39] for any strategic and operational direction in a pandemic. This requirement emerged very strongly as a theme and was highlighted at various stages, including at the team level in the ICU, at the organisational level and at the state level. Contrary to the experience in other centres [40] where there has been a significant lack of senior leadership presence at the coalface, participants in this study witnessed strong and visible leadership among ICU medical and nursing staff and at the organisational and state level. Having a strong peer-support model [41] with a flat hierarchy was appreciated by many, which resonated across both nursing and medical colleagues.

Finally, one of the significant constraints on the frontline clinicians was social distancing and the need to be physically isolated if unwell. Restrictions on mobility and gathering in public places created numerous problems in visitation hours in the ICU and in managing end-of-life situations. This experience is like other centres [42] that have dealt with managing critically ill patients with COVID-19. Resorting to technology and using video-calling resources was helpful in this space, and the lessons from this experience could be used even in non-COVID-19 situations, particularly for families from rural and remote communities. However, standard operating procedures based on established national guidelines [12] on end-of-life care, which promote space and capacity for loved ones to spend time with a dying family member, were not practised in this centre because of the highly contagious nature of COVID-19 and the restrictions that were enforced; this resonates with the experiences of other centres.[43]

The ICU staff resorted to utilising technology to assist with family conversations and physical distancing requirements. Video conferences for family meetings added another layer of complexity to management. We hope this endeavour and the lessons learned will help physicians prepare to manage similar situations in the future. Applying a multifaceted and multidisciplinary quality-improvement initiative using the PDSA (plan, do, study, act) methodology [44] would help clinician administrators and health policy managers consolidate the lessons from this study and leverage the learnings for crisis/disaster management in the future. The systems re-engineering model [45] focusses on the interaction between human factors, resource constraints and complex health systems with competing demands in managing crisis situations. While a detailed review of such interplay was outside the scope of this study, the study provided insights into some of the human factors, resource supply and use, and organisational factors. Further research on this relationship may benefit from incorporating the system re-engineering model in crisis management.

## CONCLUSION

The overall impression of frontline clinicians towards the preparedness for, and response to the COVID-19 pandemic in an ICU at this quaternary referral centre under the governance of a crisis management framework was favourable, due mainly to the enablers in ensuring wide stakeholder engagement, shared responsibilities and single point accountability.

However, we have identified opportunities for improvement in the domains of having foresight with planning and logistics (including self-sufficiency with equipment), conducting oversight of operational and strategic activity, and developing insights into communications and managing end-of-life care with compassion and with the appropriate use of technology. The line of sight of the leadership could potentially be optimised to provide opportunities to seek assistance and support when needed.

## Data Availability

https://doi.org/10.5061/dryad.rbnzs7hch.

## Acknowledgements

We would like to express our appreciation to all the critical care nurses and intensivists who participated actively in the in-depth interviews and focus group discussions. We would like to thank Dr Shagufta Perveen from the University of Adelaide for her professional assistance with transcription. We would also like to thank Dr Kathryn Zeitz, Director of Clinical Governance at the Central Adelaide Local Health Network for her support and permission to use the COSTAT matrix.

## Contributorship statement

KS was involved with the conceptualisation of the study, literature review, ethics application, conduct of the study and investigation, methodology and writing the original draft. PB was involved with the methodology, data analysis editing the manuscript. AM was involved with the writing review. AP and BR were involved with the ethics submission, methodology, data analysis and writing review. MAM was involved with literature review, methodology, data analysis and editing the manuscript.

## Competing Interests Statement

The authors report no competing interests and the authors had

- No association with commercial entities that provided support for the work reported in the submitted manuscript (the time frame for disclosure in this section of the form is the lifespan of the work being reported).
- No association with commercial entities that could be viewed as having an interest in the general area of the submitted manuscript (the time frame for disclosure in this section is the 36 months before submission of the manuscript).
- No similar financial associations involving their spouse or their children under 18 years of age.
- No Non-financial associations that may be relevant to the submitted manuscript.

Krishnaswamy Sundararajan

Peng Bi

Adriana Milazzo

Benjamin Reddi

Alexis Poole

Mohammad Afzal Mahmood

## Funding statement

**This research received no specific grant from any funding agency in the public, commercial or not-for-profit sectors’**

Krishnaswamy Sundararajan

Peng Bi

Adriana Milazzo

Benjamin Reddi

Alexis Poole

Mohammad Afzal Mahmood

## Data sharing statement

Data has been stored with the University of Adelaide repository and will be provided on request

## Ethics statement

This study involves human participants and was approved by the Central Adelaide Local Health Network (CALHN) Human Research Ethics Committee (HREC)

## References

1. Burrell AJ, Pellegrini B, Salimi F, et al. Outcomes for patients with COVID-19 admitted to Australian intensive care units during the first four months of the pandemic. Med J Aust. 2021 Jan;214(1):23–30.

2. Litton E, Bucci T, Chavan S, et al. Surge capacity of intensive care units in case of acute increase in demand caused by COVID-19 in Australia. Med J Aust. 2020 Jun;212(10):463–467.

3. Cook TM, El-Boghdadly K, McGuire B, McNarry AF, Patel A, Higgs A. Consensus guidelines for managing the airway in patients with COVID-19: Guidelines from the Difficult Airway Society, the Association of Anesthetists the Intensive Care Society, the Faculty of Intensive Care Medicine, and the Royal College of Anesthetists. Anaesthesia. 2020 Mar 27.

4. Warrillow S, Austin D, Cheung W, et al. ANZICS guiding principles for complex decision making during the COVID-19 pandemic. Crit Care Resusc. 2020 22 (2): 98–102

5. Australian Government Department of Health [internet]. Guidance on the use of personal protective equipment (PPE) in hospitals during the COVID-19 outbreak. Version 6 [cited 2020 Jun 19]. https://www.health.gov.au/resources/publications/guidanceon-the-use-of-personal-protectiveequipment-ppe-in-hospitals-duringthe-covid-19-outbreak

6. Wunsch H, Linde-Zwirble W, Harrison D, Barnato A, Rowan K, Angus D. Use of Intensive Care Service during Terminal Hospitalizations in England, and the United States. American Journal of Respiratory Critical Care Medicine 2009; 180:875–880.

7. Izcovich A, Ragusa MA, Tortosa F, Lavena Marzio MA, Agnoletti C, Bengolea A, Ceirano A, Espinosa F, Saavedra E, Sanguine V, Tassara A, Cid C, Catalano HN, Agarwal A, Foroutan F, Rada G. Prognostic factors for severity and mortality in patients infected with COVID-19: A systematic review. PLoS One. 2020 Nov 17;15(11): e0241955.

8. Huang C, Wang Y, Li X, et al. Clinical features of patients infected with 2019 novel coronavirus in Wuhan, China. Lancet. 2020; 395: 497–506

9. Wang D, Hu B, Hu C, et al. Clinical characteristics of 138 hospitalised patients with 2019 novel coronavirus-infected pneumonia in Wuhan, China. JAMA. 2020 Mar 17;323(11):1061–1069

10. Collopy BT, Williams J, Rodgers L, Campbell J, Jenner N, Andrews N. The ACHS Care Evaluation Program: a decade of achievement. Australian Council on Healthcare Standards. J Qual Clin Pract. 2000 Mar;20(1):36–41.

11. Bloomer MJ, Hutchinson AM, Botti M. End-of-life care in hospital: an audit of care against Australian national guidelines. Aust Health Rev. 2019 Oct;43(5):578–584.

12. Australian and New Zealand Intensive Care Society. The ANZICS statement on death and organ donation. Edition 4 Melbourne: ANZICS, 2019 https://www.anzics.com.au/wp-content/uploads/2020/07/ANZICS-Statement-on-Death-and-Organ-Donation-Edition-4.pdf

13. Starks H, Trinidad SB. Choose your method: a comparison of phenomenology, discourse analysis, and grounded theory. Qual Health Res. 2007 Dec;17(10):1372–80.

14. Reeves S, Albert M, Kuper A, Hodges BD. Why use theories in qualitative research? BMJ. 2008 Aug 7;337:a949.

15. Braun, V. & Clarke, V. Using thematic analysis in psychology. Qualitative Research in Psychology 2006; 3(2), 77–101

16. Krueger, Richard A. Focus groups: A practical guide for applied research (2<sup>nd</sup> ed), Thousand Oaks, CA: Sage publications 1994.

17. Neubauer, B.E., Witkop, C.T. & Varpio, L. How phenomenology can help us learn from the experiences of others. Perspect Med Educ 2019 Apr 8(2), 90–97.

18. O’Brien BC, Harris IB, Beckman TJ, et al. Standards for reporting qualitative research: a synthesis of recommendations. Acad Med 2014; 89:1245–51.

19. Spencer L, Ritchie J, Lewis J et al Quality in Qualitative Evaluation: A Framework for Assessing Research Evidence. The Cabinet Office, London 2003.

20. Hackett A, Strickland K. Using the framework approach to analyse qualitative data: a worked example. Nurs Res. 2019 Sep 21;26(2):8–13.

21. Tessema GA, Gomersall JS, Laurence CO, Mahmood MA. Healthcare providers’ perspectives on use of the national guideline for family planning services in Amhara Region, Ethiopia: a qualitative study. BMJ Open. 2019 Feb 20;9(2): e023403.

22. Tsang JLY, Ross K, Miller F, et al. Qualitative descriptive study to explore nurses’ perceptions and experience on pain, agitation, and delirium management in a community intensive care unit. BMJ Open. 2019 Apr 4;9(4): e024328.

23. http://sahealth.sa.gov.au [Internet]. Adelaide: South Australian Health; c2021 [cited 2021 Mar 23]. Public Healt; [about 2 screens] Available from:https://www.sahealth.sa.gov.au/wps/wcm/connect/public+content/sa+health+internet/public+health/disaster+preparedness+and+resilience

24. Sujan, M.A.; Huang, H.; Biggerstal, D. Trust and psychological safety as facilitators of resilient health care. In Working Across Boundaries; CRC Press: London, UK, 2019.

25. McCann, C.M.; Beddoe, E.; McCormick, et al. Resilience in the health professions: A review of the literature. Int. J. Well-Being 2013, 3, 60–81.

26. Reeves JJ, Hollandsworth HM, Torriani FJ, et al. Rapid response to COVID-19: health informatics support for outbreak management in an academic health system. J Am Med Inform Assoc. 2020 Jun 1;27(6):853–859.

27. Rubulotta F, Soliman-Aboumarie H, Filbey K, et al. Technologies to Optimise the Care of Severe COVID-19 Patients for Health Care Providers Challenged by Limited Resources. Anesth Analg. 2020 Aug;131(2):351–364.

28. Zarocostas J. How to fight an infodemic. Lancet 2020; 395:676.

29. Rathore FA, Farooq F. Information Overload and Infodemic in the COVID-19 Pandemic. J Pak Med Assoc. 2020 May;70(Suppl 3) (5): S162–S165.

30. Díaz-Guio DA, Ricardo-Zapata A, Ospina-Velez J, et al. Cognitive load, and performance of health care professionals in donning and doffing PPE before and after a simulation-based educational intervention and its implications during the COVID-19 pandemic for biosafety. Inez Med. 2020 Jun 1;28(suppl 1):111–117.

31. Yuriditsky E, Horowitz JM, Nair S, Kaufman BS. Simulation-based uptraining improves provider comfort in the management of critically ill patients with COVID-19. J Crit Care. 2020 Oct 3; 61:14–17.

32. Kleinpell R, Ferraro DM, Maves RC, et al. Coronavirus Disease 2019 Pandemic Measures: Reports from a National Survey of 9,120 ICU Clinicians. Crit Care Med. 2020 Oct;48(10): e846–e855.

33. Gandhi M, Yokoe DS, Havlir DV. Asymptomatic transmission, the Achilles’ heel of current strategies to control Covid-19. N Engl J Med 2020; 382:2158–60.

34. Lee SH, Juang YY, Su YJ, et al. Facing SARS: psychological impacts on SARS team, nurses, and psychiatric services in a Taiwan general hospital. Gen Hosp Psychiatry. Sep-Oct 2005; 27:352–358.

35. Fusaroli P, Balena S, Lisotti A. On the death of 100l+lItalian doctors from COVID-19. Infection. 2020 Oct;48(5):803–804.

36. Tabah A, Ramanan M, Laupland KB et al. PPE-SAFE contributors. Personal protective equipment and intensive care unit healthcare worker safety in the COVID-19 era (PPE-SAFE): An international survey. J Crit Care. 2020 Oct; 59:70–75.

37. Rangachari P, L Woods J. Preserving Organizational Resilience, Patient Safety, and Staff Retention during COVID-19 Requires a Holistic Consideration of the Psychological Safety of Healthcare Workers. Int J Environ Res Public Health. 2020 Jun 15;17(12):4267.

38. Albott CS, Wozniak JR, McGlinch BP, Wall MH, Gold BS, Vinogradov S. Battle Buddies: Rapid Deployment of a Psychological Resilience Intervention for Health Care Workers During the COVID-19 Pandemic. Anesth Analg. 2020 Jul;131(1):43–54

39. Everly GS, Wu AW, Crumpsty-Fowler CJ, Dang D, Potash JB. Leadership principles to decrease psychological casualties in COVID-19 and other disasters of uncertainty. Disaster Med Public Health Prep. 2020 Oct 22:1–10

40. I am A Nurse in A Covid-19 Unit. My Hospital’s Leaders Frighten Me More Than the Virus. Available online: https://www.statnews.com/2020/05/06/nurse-frightened-hospital-administrators-more-than-covid-19/ (accessed on 12 December 2020).

41. Adams JG, Walls RM. Supporting the Health Care Workforce During the COVID-19 Global Epidemic. JAMA. 2020;323(15):1439–1440

42. Negro A, Mucci M, Beccaria P, et al. Introducing the Video call to facilitate the communication between health care providers and families of patients in the intensive care unit during COVID-19 pandemic. Intensive Crit Care Nurs. 2020 Oct; 60:102893.

43. Sasangohar F, Dhala A, Zheng F, Ahmadi N, Kash B, Masud F. Use of telecritical care for family visitation to ICU during the COVID-19 pandemic: an interview study and sentiment analysis. BMJ Qual Saf. 2020 Oct 7: bmjqs-2020-011604.

44. Taylor MJ, McNicholas C, Nicolay C, Darzi A, Bell D, Reed JE. Systematic review of the application of the plan-do-study-act method to improve quality in healthcare. BMJ Qual Saf. 2014 Apr;23(4):290–8.

45. Holden RJ, Carayon P, Gurses AP, Hoonakker P, Hundt AS, Ozok AA, Rivera-Rodriguez AJ. SEIPS 2.0: a human factors framework for studying and improving the work of healthcare professionals and patients. Ergonomics. 2013 Nov 1;56(11):1669–86.

